# Mapping Gene-by-Environment Interactions via the Plasma Exposome at Cell-Type Resolution

**DOI:** 10.64898/2026.11.16.26368215

**Authors:** Haim Krupkin, Marie Amale Huynh, Daniel Nachun, Peter Durda, Yongmei Liu, Kent D. Taylor, Robert E. Gerszten, Clary B. Clish, François Aguet, Kristin Ardlie, W. Craig Johnson, Russell P. Tracy, Stephen S. Rich, Jerome I. Rotter, Stephen B. Montgomery

**Affiliations:** Department of Genetics, Stanford University, Stanford, CA, 94305, USA; Department of Pathology, Stanford University, Stanford, CA, 94305, USA; Department of Biomedical Data Science, Stanford University, Stanford, CA, 94305, USA; Department of Pathology and Laboratory Medicine, LArner College of Medicine, University of Vermont, Burlington, VT, 05446, USA; Divisions of Cardiology, Duke University Medical Center, Durham, NC, 27710, USA; The Institute for Translational Genomics and Population Sciences, Department of Pediatrics, The Lundquist Institute for Biomedical Innovation at Harbor-UCLA Medical Center, Torrance, CA, 90502, USA; Department of Public Health Genomics, Beth Israel Deaconess Medical Center, Boston, Boston, MA, 02215, USA; Broad Institute, MIT and Harvard, Cambridge, MA, 02215, USA; Department of Biostatistics, University of Washington, Seattle, WA, 98195, USA; Department of Pathology and Laboratory Medicine, Larner College of Medicine, University of Vermont, Burlington, VT, 05405, USA; Division of Cardiovascular Medicine, University of Virginia School of Medicine, Charlottesville, VA, 22908, USA.

**Author notes:** Corresponding author: Stephen B. Montgomery,. Deceased (Russell P. Tracy).

**Keywords:** Quantitative Trait Loci, Multi-Omics, Exposome, Gene-by-Environment, Systems biology

## Abstract

Gene-by-environment (GxE) interactions inform where genetics and environment converge to impact human health and disease. However, systematically mapping GxE interactions has been challenging due to the diversity of testable exposures and a lack of scalable approaches to measure the environments cells experience *in vivo*. Here, we identified modules of correlated plasma metabolites and proteins and used them as quantitative proxies for environmental exposures experienced by circulating leukocytes. We demonstrated that these proxies are associated with environmentally-modifiable clinical traits such as diet and infection and support broad mapping of gene-by-environment (***G × E***) interactions affecting gene expression and CpG methylation in leukocytes. We identified significant interactions for 95.2% of tested genes and 5.9% of tested methylation sites. These ***G × E*** interactions showed specificity to exposures and cell type. Colocalization analysis further provided links between interactions and complex trait and disease loci. Finally, we expanded ***G × E*** interaction mapping to unexplored cell types using a zero-shot transfer model to predict ***G×E*** interactions in held-out cell types, demonstrating how plasma metabolomics and proteomics can enhance the discovery of ***G × E*** interactions across the body. Altogether, this work provides a systematic multi-omic framework to study how gene regulation is shaped by both the genome and the molecular environments cells experience *in vivo*.

## 1 Introduction

Genome-wide association studies (GWAS) enable identifying genetic variants linked to complex traits and disease. However, GWASes often do not account for environmental contexts and can miss genetic variants whose effects are modified by the environment. Investigating these *G × E* interactions has been challenging due to a several reasons. Traditional *G×E* studies typically test for interactions with predefined environmental variables, such as self-reported diet, smoking status, and exercise-induced changes^1–5^. In addition, detecting a *G × E* interaction requires approximately four times the sample size needed to find a direct genetic or environmental effect of the same strength, increasing the cost of such studies^6^. Furthermore, these approaches are often restricted in scope to a few cell types or environments, limiting their ability to systematically capture *G × E* interactions across the body.

One opportunity to address the bias of pre-defined environmental variables lies in using blood plasma itself, as the variance in the blood plasma metabolome and proteome is largely influenced by environmental factors^7–9^. For instance, although variation in blood carbohydrate levels is partially genetically determined, it is mostly driven by environmental exposures such as meal timing.

Furthermore, studying gene-by-environment (*G×E*) interactions effects on molecular phenotypes such as gene expression and methylation has previously demonstrated improved power to identify variants where genetics and specific environments converge to affect gene regulation.^10–12^.

We take advantage of both of these opportunities and use the axes of variation in the plasma metabolome and proteome as environmental proxies for the discovery of *G × E* quantitative trait loci for molecular phenotypes (gene expression and methylation). Using multi-omic data from the Multi-Ethnic Study of Atherosclerosis (MESA)^13^, we derive co-varying modules of blood plasma metabolites and proteins (which we refer to as “Molecular Exposure Modules”, or MEMs) and use them as proxies for environmental states. We propose that these major axes of variation in the plasma metabolome and proteome represent the major exposures experienced by individuals. By quantifying interactions between these MEMs and the genome, we map *G × E* effects on cell type specific gene expression and DNA methylation and identify pervasive interaction effects. We then link interactions to complex traits through colocalization, bridging genetic variants, molecular environments, gene regulation, and complex traits. This approach provides a comprehensive multi-cell-type landscape of *G × E* effects across the leukocyte transcriptome and methylome.

## 2 Results

### 2.1 Axes of molecular variance in blood plasma

To systematically map *G× E* interactions, we developed an integrated multi-omic workflow utilizing data obtained from MESA (Fig. 1A). We chose to focus our analysis on the data from participants with complete multi-omic profiles(N=541; Fig. 1B; see Methods 4.1). When samples were collected, blood plasma and blood leukocytes were separated; blood plasma was retained for metabolomic and proteomic profiling (see Methods 4.2), and blood leukocytes was retained for transcriptomic and methylomic profiling^12^.

To study the main axes of variation of the metabolome and proteome, we performed a weighted gene co-expression network analysis (WGCNA) on the proteome and metabolome (see Methods 4.3). We refer to the resulting modules as MEMs, as they primarily represent the molecular exposure^7–9^ to which blood cells are exposed. The WGCNA analysis resulted in a total of 52 metabolic MEMs, and 18 proteomic MEMs. WGCNA links metabolites/proteins to a single MEM, thus enabling a high degree of explainability for each MEM. Furthermore, we can characterize each MEM by its components, testing to see if a MEM’s components are enriched in a specific annotation (see Methods 4.4; Extended Fig. 1; supplementary tables 1 and 2). On average, each metabolomic MEMs captures 51% of the percent variance explained (PVE) within its module, and combined they explain 37.1% of the PVE across the entire metabolome. Each proteomic MEMs captures on average 11.8% of the PVE within its module, and a total 10% of the PVE across the entire proteome (Fig. 1C; see Methods 4.5). To test for cross-ome redundancy, we performed pairwise correlations, finding no pair of MEMs has a meaningful correlation (*R*^2^ *>* 0.01), indicating that MEMs represent distinct information (Extended Data Fig. 2).

**Fig. 1:**
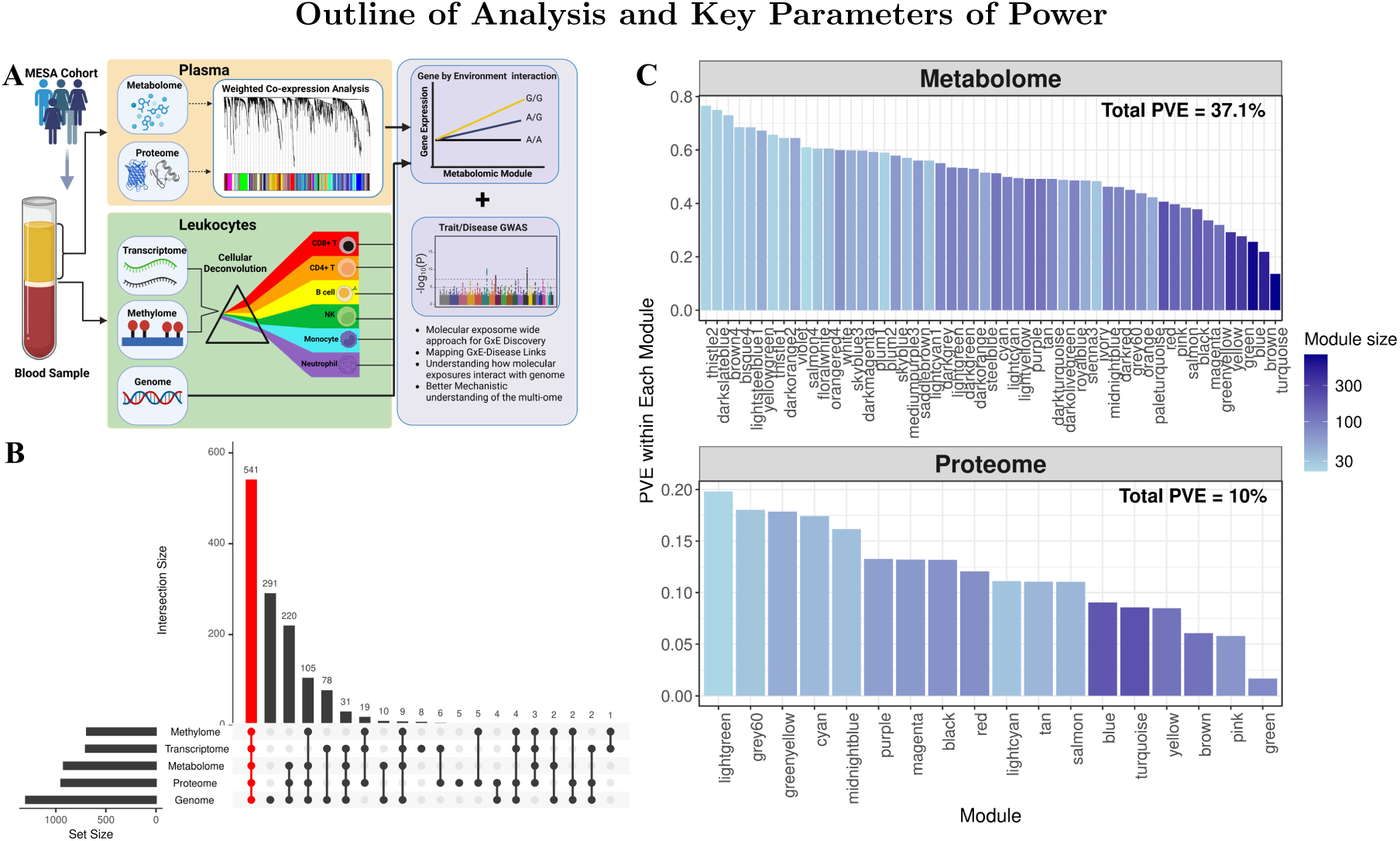
**A**, Schematic of workflow. We start with samples of blood leukocytes and plasma. The plasma is profiled via proteomics and metabolomics, the leukocytes are profiled via transcriptomics and methylomics. The bulk transcriptomic and methylomics are de-convoluted to be cell type specific. In parallel, the axes of variation in the proteomics and metabolomics are identified via WGCNA, into Molecular Exposure Modules (MEMs). Because the metabolomics and proteomics are measured in the plasma, wherein the leukocytes largely reside, they provide a window into the molecular environment the cells are exposed to, enabling unsupervised study into how the genome of a cell interacts with its environment. **B**, Upset plot representing the number of individuals for whom we have collected a samples of each ome. **C**, PVE of each proteomic and metabolomic MEM (see Methods 4.5).

Having identified MEMs, we tested whether they captured meaningful environmental information. We found that many of the 70 MEMs had strong correlations with environmentally-modifiable clinical phenotypes (Methods 4.6, Supplemental Table 3). In particular, some metabolomic MEMs were found to be strongly associated with lipid traits, for example, with LDL, HDL, and triglyceride levels. Additionally, some immune traits show strong associations with proteomic MEMs, particularly IL-6 and C-Reactive protein (CRP). Proteomic MEMs also correlated with BMI and weight, su pporting the established link between BMI and inflammation^14,15^. Similarly, a group of clustered metabolic MEMs were associated with blood triglyceride and cholesterol levels (Fig. 2).

**Fig. 2:**
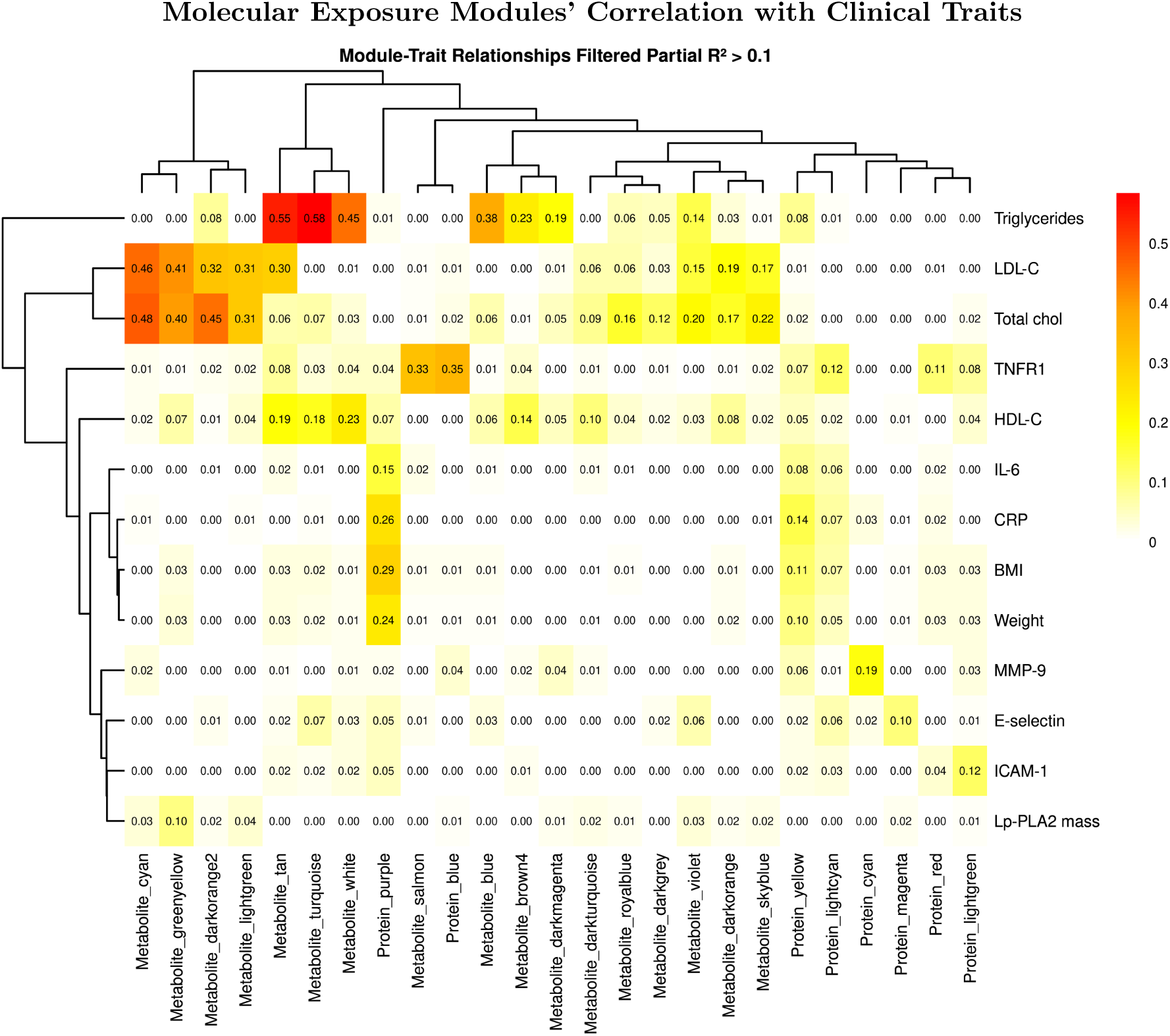
Associating MEMs with clinical traits via multi-variate linear regression with sex, self-identified ancestry, measurement site, and age of measurement as covariates to reduce confounding. Displayed are a subset of MEMs and clinical trait for which there is at least one pair with high variance (*R*^2^ *>* 0.1; see Methods 4.6).

### 2.2 Discovery of Gene by Molecular Exposure interactions

We tested whether genetic variants associated with altered gene expression and methylation were interacting with molecular exposure defined by MEMs. We employed a *G × E* analysis, utilizing MEMs as the environment (see Methods 4.8). Applying this approach, out of the 16,750 tested genes, we identified 15,953 (95.2%) unique eGenes with a *G × E* interaction. For CpG methylation sites, out of a total 159,161 tested CpG sites, 9,472 unique *G × E* CpG sites (5.9%) harbored a *G × E* interaction. This analysis was performed for each cell type, across all MEMs. Overall, this rate of discovery is similar to what was previously reported: previous evidence shows that 32.2% of genes exhibit *G × E* interactions^11^, and 3.2% of methylation sites^16^.

Importantly, our higher discovery rate might be attributable to the higher scale and statistical power of our analyses. Specifically, we analyzed data from more individuals than the prior studies (541 versus 249 for eGenes, and 149 for methylation sites). We also examined a broader range of cell types (6 versus 1) as well as environments (70 versus 4). Together, all these factors likely increased our ability to detect significant associations. Overall, these findings suggest that *G × E* effects on gene expression are observed widely. To examine the validity of the observed *G × E* interaction, we evaluated the overlap with a previously reported set of *G × E* interaction eQTLs, related to immune activation^11^. We found an enrichment of overleaping *G × E* interactions (odds ratio=1.33, *p.value <* 0.001). Although the environmental context is different, this overlap further validates the observed association *G × E* interactions.

We observed a wide heterogeneity in the forms of *G×E* interactions, with some variants exhibiting only a main genetic effect (Fig. 3A left), some exhibiting only a *G × E* interaction (Fig. 3A middle), and some exhibiting both (Fig. 3A right). For example, CD36 gene expression in monocytes shows a *G × E* interaction with metabolic MEM Tan (Tan is correlated with the clinical traits: Triglycerides, LDL, HDL; Fig. 2). This recapitulates CD36’s known environment dependent role as a lipid scavenger receptor^17^, suggesting *G × E* variants alter monocytes’ response to circulating lipid levels (Fig. 3A center). For each individual MEM, many associated molecular phenotypes were identified. For example, for a randomly selected cell type-MEM-molecular phenotype combination; Monocyte-Metabolic MEM Yellow-expression; we found *G × E* interactions across the genome (Fig. 3B). Of note, this analysis is observational, and thus does not determine causality.

**Fig. 3:**
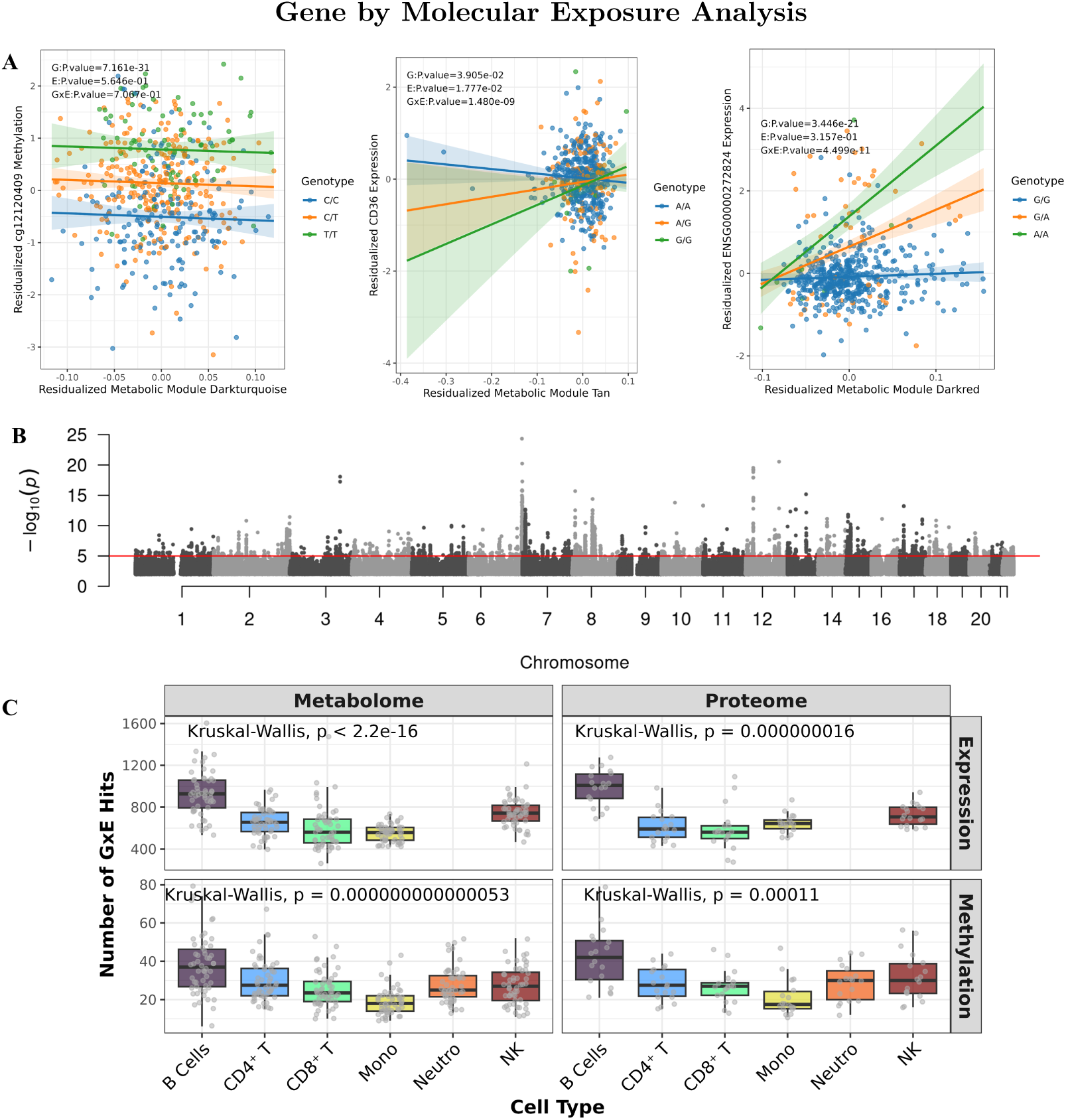
**A**, Examples of variants, and their association with molecular phenotypes, and whether a MEM is associated with a change in molecular phenotype levels. Left panel shows a main effect methylation affecting variant *chr6:30444190 C > T* associated with altered CpG methylation levels of cite cg12120409, and is independent of the affects of the metabolic MEM Darkturquoise. Middle panel shows a *G×E* interaction variant *chr6:30444190 T > A* associated with altered CD36 expression, and is interacting with metabolic MEM Tan. Right panel shows both a main effect and a *G×E* interaction between the variant *chr1:148438355 G > A*, the metabolic MEM Darkred, and the gene ENSG00000272824. **B**, Example manhattan plot of the monocyte expression associated *G×E* interaction variants with metabolic MEM Tan. **C**, Number of *G×E* molecular phenotype interactions by cell type. * Also, of note, although some module names are shared between the metabolome and proteome, they do not represent the same axes of variation, nor are they composed of the same components.

Next, we examined the observed heterogeneity in the number of *G×E* hits across MEMs, molecular phenotypes, omes, and cell types (Fig. 3C; Fig. 4A), and asked which factors explain this variation (see Methods 4.9). We found that the strongest determinant of the number of *G×E* hits was the number of main effect hits. The next strongest determinant was cell type, with B cells the highest number of *G×E* associated molecular phenotypes, adjusted for covariates, suggesting they are the most environmentally responsive . In contrast, the topological features of the MEM, such as the module size, density, and coreness in the overall metabolome/proteome network, explained little of the variance in the number of *G×E* interactions with a specific MEM (Extended Fig. 3A). We next examined the universality of *G × E* interactions. Many molecular phenotype associated with a *G × E* interaction were unique to a single cell type, with this effect being even more evident among methylation *G × E* interactions (Fig. 4B), than with *G×E* interactions affecting gene expression. This is consistent with previous observations that methylation is more cell type specific than gene expression^18^. To better understand the principles governing which molecular phenotypes have a *G × E* interaction with a MEM, we compared the molecular phenotypes identified in each cell type-MEM-ome combination. Generally, molecular phenotypes were specific to an individual MEM, but nonetheless, some were highly shared (Extended data Fig. 4A). We found that eGenes were generally more shared across cell types than methylation sites. In addition, we observed molecular phenotypes associated with a *G × E* interactions were more similar across cell types than across omes (Fig. 4B-C).

Next, we sought to understand molecular phenotypes with *G × E* interactions across multiple MEMs. We examined the pairwise correlations among the number of proteomic MEMs, the number of metabolomic MEMs, and the number of cell types in which a given molecular phenotype is associated with a MEM. We find a consistent additive pattern. Specifically, molecular phenotypes that harbor a *G × E* interaction tend to span more MEMs across multiple omes, and this cross-omeness further correlates with the number of cell types in whom the molecular phenotype harbors a *G×E* interaction. This effect is especially evident for eGenes, but was also found to a lesser degree in methylation, possibly due to the cell specific nature of methylation (Extended data Fig. 4B; see Methods 4.10).

To address the limited scalability of *G × E* mapping across many cell types, we asked whether interaction patterns learned in observed cell types could be transferred to a held-out cell type using a machine learning approach. As an initial, proof of concept framework for zero-shot transfer learning of *G × E* interactions between cell types using gene expression or methylation as the input features, we trained an XGBoost tree model. The model was trained to learn, based on the consistency of the molecular phenotype’s *G × E* interaction in the observed cell types, as well as the cell types expression pattern, whether in a held-out cell type there will be a *G × E* interaction. Across cell types, the model performed better than random, achieving a lift of 2.11-3.3 for expression, and a lift of 2.3-3.4 for methylation (Fig. 4D; see Methods 4.11).

To shed some light into the inner workings of the model, in particular the eGene *G × E* prediction model as an exemplar, we used SHapley Additive exPlanations (SHAP)^19^ to determine feature importance for predicting the presence of an eGene-MEM pair. We found gene responsiveness (gene_resp_rank), defined as the rank of gene according to how many unique MEMs harbor a *G × E* with the gene, was the most important feature. We also found the strength of the MEM (log_module_potency_gene_n_nodules) to also be important to determining whether a *G×E* interaction was detected (Extended Data Fig. 5).

**Fig. 4:**
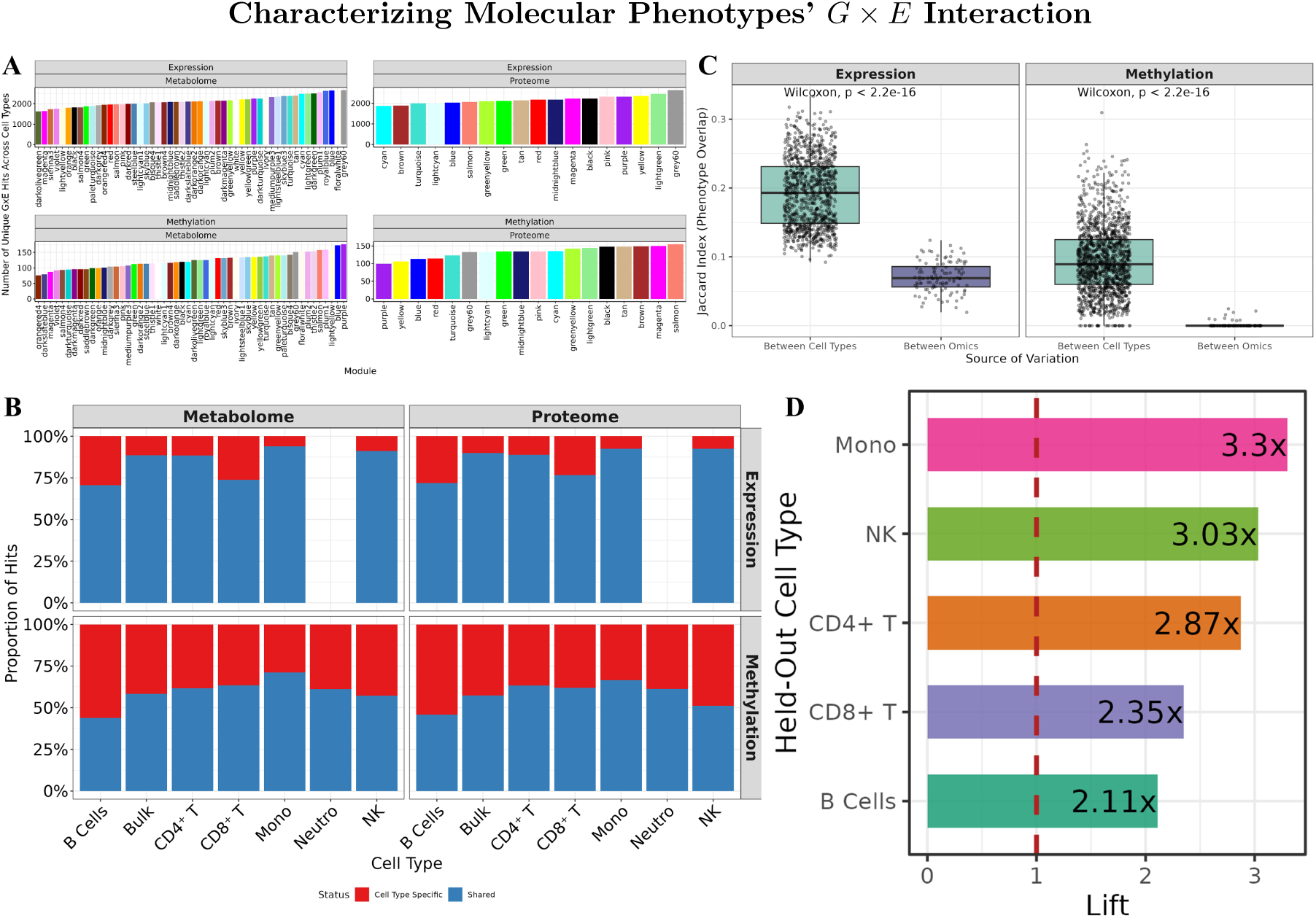
**A**, Number of unique eGenes or CpG sites for whom *G × E* interaction is found in monocytes, across proteomic and metabolic MEMs. The y axis is not constant, as there are more eGenes then there are methylation sites for which the level is altered as a function of a *G × E* Interaction. **B**, Proportion of *G × E* molecular phenotypes who are shared across more then one cell type, or that are found exclusively in that cell type. **C**, Comparison of the molecular phenotypes found to be associated with each MEM via a *G × E* interaction, comparing across cell types, and across omes. **D**, Lift - Fold change compared to random, of the Expression-MEM *G × E* prediction using a simple XGBoost tree models model predicting whether for a left out cell type a molecular phenotype will have a *G × E* interaction in a specific MEM (see Methods 4.11).

### 2.3 Genetically Associating Molecular Exposure Modules with Complex Traits

Following our analysis of the factors governing *G × E* interactions, we sought to understand whether these interactions were relevant to human traits, especially diseases. To explore this goal, we performed colocalization analysis between human genome wide association studies (GWAS)^20^ and the *G×E* interactions identified here (see Methods 4.12). We observed colocalizations across a wide array of complex traits (supplementary table 4), some of which did not colocalize at the main effect QTL layer. For example, we observe a colocalization between MCAM in CD8^+^ T cells, metabolic MEM Mediumpurple3, and Rheumatoid arthritis (RA; Fig. 5A). MCAM positive CD8^+^ T cells are known to be associated with autoimmune disease, and in particular RA^21^. They are enriched in the synovial fluid of RA patients, and have a known role in secreting the pro-inflammatory cytokine IL-17. Moreover, MCAM is also thought to facilitate the migration of MCAM positive CD^+^ T cells into RA patient’s inflamed joint tissue. Furthermore, the metabolic MEM MediumPurple3’s 10 most strongly associated clinical traits, ordered by variance explained, wer:; E-selectin (*R*^2^ = 0.078), TNFR1, BMI, weight, antihypertensive medications, triglycerides, IL-6, CRP, LpPLA2 mass, and RBC count (*R*^2^ = 0.014). This group of clinical traits is consistent with the well-established association between metabolic syndrome and RA risk^22^. Overall, these results hint a possible *G × E* interaction affecting MCAM expression in CD8^+^ T cells within a metabolic-inflammatory environment, with a suggested role in RA. Another example of *G × E* colocalization was observed in the bulk analysis, involving AEBP1 and the metabolic MEM Magenta, and high cholesterol (Fig. 5B). AEBP1 is a known transcriptional repressor of the genes enabling removal of cholesterol^23^. In general, we are able to find many *G × E* interactions that colocalize with GWAS hits, with a median percentage of 68.8% (CI: 61.9-75.6%) of all molecular phenotype-GWAS pairs being *G × E* interactions (Fig. 5C).

**Fig. 5:**
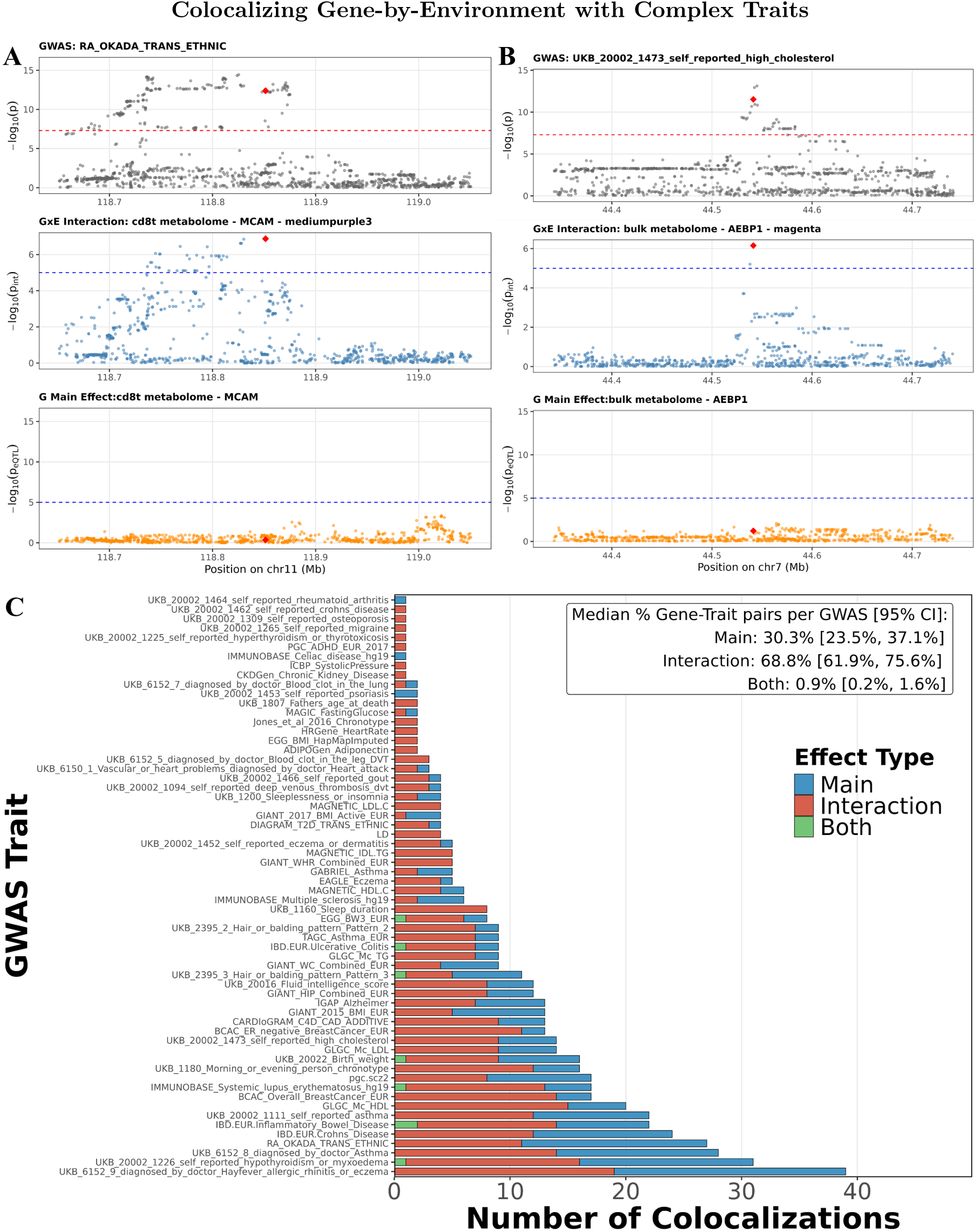
Specific examples of colocalizations between complex trait GWAS and *G × E* interactions compared to main effects. **A**, Example locus on chromosome 11 wherein a colocalization is found between a RA GWAS and a *G × E* interaction with the metabolic MEM MediumPurple3, associated with MCAM gene expression levels in CD8^+^ T cells **B**, Example locus on chromosome 7 wherein a colocalization is found between a high cholesterol GWAS and a *G × E* interaction with the metabolic MEM magenta, associated with AEBP1 gene expression levels in the bulk analysis. **C** Number of unique colocalized molecular phenotype-GWAS pairs of each type, of each type; main effect, *G × E* interaction, and pairs discovered in both.

## 3 Discussion

Efforts to map *G × E* interactions have largely relied on either *in vitro* stimulation, for example by exposing cell lines to immune modulators^11,24,25^ or on mapping interactions with broad epidemiological variables such as age, BMI, and diet^26–28^. The first offers causal mechanistic clarity but limited scope; the second captures real-world tangible variation but lacks explainability and molecular resolution. A third approach, testing interactions with individual metabolites or proteins across the entire metabolome/proteome would be prohibitive due to the correlated structure of each measurement and multiple testing. None of the approaches mentioned provides a scalable framework to systematically connect the molecular environment a cell experiences *in vivo* to its molecular phenotype.

Thus, in this work, we took a complementary approach. We systematically quantified *G × E* interactions across omic-layers using the plasma metabolome and proteome as quantitative proxies for the molecular exposures that blood leukocytes are exposed to *in vivo*. We constructed our quantitative proxies for molecular exposures in an unsupervised manner via network analysis using WGCNA, and called them Molecular Exposure Modules (MEMs). MEMs provide a framework to simplify the metabolome and proteome, offering a degree of explainability, as each metabolite/protein is assigned exclusively to a specific MEM, which allows a later over representation analysis to annotate each MEM. For example, we found MEMs associated with fatty acid metabolism and inflammation.

Next, we tested the effects of genotype-by-MEM interactions on cell type specific gene expression and methylation across 541 individuals from MESA. Characterization of the *G × E* interactions revealed that *G × E* are not centralized to a specific gene, CpG site, cell type, or MEM. Rather, the data indicates that *G × E* interactions are pervasive, occupying a spectrum of specificity, with methylation *G × E* interactions tending to be more cell-type specific. Moreover, an additive pattern emerges: molecular phenotypes (either gene expression or CpG methylation) for which a *G × E* interaction is found with multiple MEMs, are observed to be associated with multiple proteomic and metabolomic MEMs, and with multiple cell types. However, the strongest predictor of whether a gene will harbor a *G × E* interaction is simply whether a genomic main effect is discovered for that gene in that cell type. Among all cell types examined, B cells were found to have the highest number of detected unique molecular phenotypes per context, both with and without adjustment for potential covariates, and despite their limited proportion in the samples. This suggests that B cells are either the most environmentally responsive cell type, or alternatively, that a systematic bias is inflating false positives in B cells. Overall, these observations improve our understanding of the governing rules of context-dependent genetic regulation.

Currently, profiling *G×E* interactions across all cell types and molecular exposures is prohibitive for many reasons, including computational complexity, limited statistical power, and cost. We sought to demonstrate that the structure of *G × E* interactions is not random but learnable. By utilizing a zero-shot transfer learning framework trained on *G × E* interaction patterns, we show *G × E* interactions are predictable in a held-out cell. This provides a path towards scaling up the discovery of *G × E* interactions across the body when plasma proteomics and metabolomics are collected alongside multi-tissue multi-omics data. Further, as the number of cell types with characterized molecular profiles expands with the rise of single cell technologies^29–32^, together with the increase in proteomic/metabolomic profiling^33^, predictive frameworks may enable the scaling of *G× E* mapping into new cell types.

As mapping molecular *G × E* interactions has enabled the discovery of “missing regulation” for complex traits and diseases, we also demonstrate how our approach can systematically uncover *G×E* effects contributing to health and disease. Colocalization with complex trait GWAS signals indicated that several of our *G × E* interactions may contribute to human phenotypes. Across a range of complex traits, we saw a median of 68.8% of colocalizations due to *G × E* interactions. We see this work enabling more systematic linking of molecular environment-dependent molecular regulation to disease-relevant genetic architecture.

This work has several limitations. Regarding statistical power and design, our sample size (N=541), while large relative to comparable studies^6^, remains modest for *G×E* mapping. Our analysis is also cross-sectional and regresses out the effects of sex and ancestry. Further, current technologies do not capture all metabolites and proteins. Moreover, we do not account for heterogeneity in the biological importance of different metabolites and proteins, and we are limited in the scope of cell type deconvolution to abundant leukocytes. Regarding methodological limitations, we expect that further adaptations of our zero-shot prediction model may benefit from integrating prior knowledge, such as known eQTLs, regulatory annotations, or external databases. Furthermore, the colocalization procedure we employed is conservative, as we did not use variant bayesian fine-mapping^34^ nor account for cross-MEM pleiotropy^35^. Moreover, as with many *G × E* studies, we do not establish causal directionality between the environment (MEMs) and molecular phenotypes, although MEMs might be a heritable trait of blood leukocytes. This is similar to *G × E*, in which BMI is modeled as the environment, although BMI has high heritability^36,37^.

In conclusion, we present a map of gene-by-environment interactions with the plasma molecular environment, revealing widespread context-dependent genetic regulation. Constituting one of the most comprehensive multi-omic *G × E* mapping efforts to date, this work is a step towards understanding how the genome, and molecular exposures affect phenotype.

## 4 Methods

### 4.1 The MESA Multi-Omic Cohort

The Multi-Ethnic Study of Atherosclerosis (MESA) is a population-based cohort that is part of the Trans-Omics for Precision Medicine (TOPMED) project. Specifically, MESA aimed to integrate whole-genome sequencing with multi-omic and clinical data to advance precision medicine. As part of this effort, MESA has longitudinally collected biological and clinical measurements from diverse participants from across the U.S. For the present analysis, we focused on the MESA multi-omic pilot data. In particular, the first of two examination periods conducted approximately ten years apart: Exam 1 (July 2000 to August 2002) and Exam 5 (April 2010 to February 2012). Generally, MESA is a study of the characteristics of subclinical cardiovascular disease and the risk factors that predict progression to clinically overt cardiovascular disease or progression of the subclinical disease^13^. Furthermore, the general MESA cohort consists of a diverse, community-based sample of an initial 6,814 men and women aged 45-84 years without known cardiovascular disease at baseline. Thirty-eight percent of the recruited participants were White, 28 percent African American, 22 percent Hispanic, and 12 percent of Chinese descent. Participants were recruited from six field centers across the United States: Baltimore City and Baltimore County, Maryland; Chicago, Illinois; Forsyth County, North Carolina; Los Angeles County, California; New York, New York; and St. Paul, Minnesota.

We chose to focus only on participants who have both exam 1 and 5 data. As part of the pilot study, a subset of participants was profiled using a multi-omic framework, including plasma metabolomics (6,052 metabolites) and proteomics (1,317 proteins), as well as RNA sequencing and DNA methylation measurement to capture gene expression and methylome epigenetic variation. The samples from MESA participants were selected for the TOPMed multi-omic pilot study due to the availability of previously collected specimens and the high ancestry diversity of the cohort. In addition, genotype data is available for all individuals in the multi-omic study. After harmonizing and intersecting samples across all molecular modalities and both exams, a total of 541 participants had complete data across all omes at both time points, forming a deeply phenotyped cohort, used for an integrative multi-omic study of the genome and molecular exposome^13,38^. For a detailed outline of methods and quality control for MESA, refer to the supplemental methods section in^12^.

The MESA study was approved by the Institutional Review Boards at all participating institutions, and all participants gave written informed consent. In addition, informed consent was obtained for extensive data sharing (dbGaP) and genetic/omic studies, including candidate genes (NHLBI CARe), genome-wide scans (NHLBI SHARe), exome sequencing (NHLBI ESP).

### 4.2 Metabolome Annotation

Metabolomic profiles for MESA samples were created by the Broad Institute using and liquid chromatography–mass spectrometry (LC-MS). Detailed experimental protocols have been reported elsewhere^39–43^.

We processed the initial dataset of 6,042 detected metabolite LC-MS peaks. Initially, annotations of 492 (8.1%) were made using reference standards. To predict additional IDs, we employed MetID^44^, an R package specifically designed for metabolite identification. This tool utilizes accurate mass-to-charge ratios (m/z) and retention times (RT) to putatively identify unknown metabolites. We leveraged seven publicly available databases: HMDB^45^, KEGG^46^, MassBank^47^, NIST, T3DB^48^, BloodExposome^49^, and DrugBank^50^. The best annotation match for each metabolite was retained based on the highest confidence from the available databases. As a result, out of the 5,962 initially unannotated metabolites, 5,938 (99.6%) were putatively annotated, representing a significant increase in coverage.

Of the original 6,052 detected metabolites, we putatively matched 3,085 unique compounds, suggesting a substantial level of duplication (2,856 metabolites, or 48.3%). This redundancy likely arises from mapping LC-MS peaks from isomeric compounds to a single database compound and the lack of retention time (RT) data in the databases used for mapping. Further, since RT is not considered in the matching process, multiple annotations could be assigned to the same metabolite LC-MS peak based solely on mass-to-charge ratios. A limitation to note is that RT varies greatly depending on the chromatic conditions, making retention times across different labs difficult to compare. Finally, while the three nontargeted LC-MS methods used to acquire the data employed different chromatography methods (hydrophilic interaction liquid chromatography (HILIC)-positive ion MS for polar metabolites, reversed phase C8 chromatography-positive ion MS for lipids, and reversed phase C18 chromatography-negative ion MS for compounds of intermediate polarity), there is overlapping coverage of subsets of metabolites. Thus, it is ultimately important to confirm identities of significant metabolite features using authentic reference standards.

### 4.3 Mapping Molecular Exposure Modules

To map the molecular exposome by using the metabolome and proteome as proxies, we used Weighted Gene Co-expression Network Analysis (WGCNA)^51^. WGCNA is a method aimed at building a biologically meaningful network to infer relationships between metabolites or proteins. In this context, “gene” co-expression networks can help us construct modules of co regulated metabolites/proteins and thus gives us biologically inferable modules of metabolites that act similarly. For example, if glucose and another carbohydrate are correlated, they should be in the same module.

One advantage of using WGCNA is that we do not need to specify the number of modules in advance, reducing bias. However, some bias remains, because researchers still indirectly choose parameters that affect the number and composition of the modules. We follow the pipeline summarized in Extended Data Fig. 6 to chose the parameters for WGCNA. For normalization, we explored both rank norm normalization and variance stabilizing transformation (VST). After examining the relationship between the rank of the mean values and the standard deviation (SD) of metabolites and proteins across both exams, we selected VST normalization as it effectively reduces variability across ranks, ensuring more consistent variance for downstream analyses. We adjusted for both ancestry PCs and batch effects. Specifically, we removed the first principal component, as well as ancestry PCs (Extended Data Fig. 7).

### 4.4 Annotating Molecular Exposure Modules

To characterize the metabolic MEMs, we performed over-representation analysis (ORA) against the Human Metabolome Database (HMDB)^45^, specifically chemical information found in the Classy-Fire fields, deciphering what chemical classes are enriched within each metabolic MEM. To ensure robust results, annotation terms were filtered to include only those containing between 3 and 500 metabolites. To characterize the proteomic MEMs, we performed ORA using the enrichR package^52^, specifically against Gene Ontology^53^, Reactome^54^, and KEGG^46^. Nominal *p*-values from both enrichment analyses, were separately adjusted for multiple testing using the Benjamini-Hochberg False Discovery Rate (FDR) method. A significant hit was defined as a hit with an *FDR <* 0.05.

### 4.5 Calculation of Molecular Exposure Module Variance Explained

To evaluate the representational power of the MEMs, we evaluated the proportion of variance explained (PVE) by all the MEM’s, as well as for each individual MEM. Specifically, to measure the proportion of variance explained (PVE) within each individual MEM, we used the “`propVarExplained`” function from the WGCNA package^51^. To calculate the total variance explained across all MEMs for a specific ome(*PV E_T otal_*), we used the following equation:

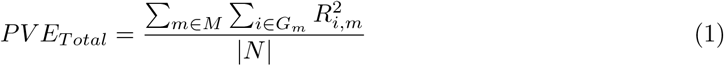

where *M* is the set of all identified MEMs (excluding the unassigned features-grey MEM), *G_m_* is the set of features assigned to module 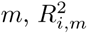 is the squared Pearson correlation coefficent between gene *i* and its module’s MEM, similarly to the PVE metric computed internally by WGCNA per MEM and *|N|* is the total number of features in the dataset (including the unassigned grey features).

### 4.6 Associating Molecular Exposure Modules With Traits

To handle possible hidden confounders, we performed an multi-variate regression analysis, wherein sex, self-identified ancestry, site, age and age at measurement were included as covariate to isolate the specific association between each clinical trait and module score. Statistical significance was defined as *p <* 0.05. Our primary focus was the variance explained (*R*^2^) by each association.

### 4.7 Methylome and Transcriptome Pre-processing and Cell Type De-convolution

Generally, we pre-processed the transcriptome and methylome of samples, we then deconvolved the bulk data into cell type specific data. In principle, this is similar to what was done previously^12^.

#### Transcriptome

To estimate the cell-type-specific gene expression profiles from the bulk MESA RNA-sequencing data, we utilized Tensor Composition Analysis (TCA)^55^. Specifically, we first took the bulk expression and normalized it to counts per million (CPM). To mitigate noise and focus on biologically relevant signals, we performed feature selection to identify highly variable genes (HVGs) using the scran R package^56^. We retained only the consensus HVGs that appeared in the top highly variable sets. Then, we took this bulk RNA-sequencing data, and using CIBER-SORT^57^ we estimated the prevalence of each cell type using the default leukocyte reference matrix. We then excluded cell types with less than a median of 0.1% proportion. Then, using the afore-mentioned proportions of each cell type, we utilized TCA to retrieve cell type specific expression profiles. Then, to enable downstream account for unobserved confounding variation (e.g. batch or population structure affects), we performed PCA, and approximated the optimal number of principal components as covariances by using the Gavish-Donoho method^58^. Transcriptomics were done on the PBMCs.

#### Methylome

Generally, we followed a similar pipeline to the one we employed on the transcriptome. Specifically, we first pre-processed the data, then we estimated cell type proportions from the bulk methylation data, then we used TCA to de-convolve cell type specific individual methylome profiles. In particular, we first employed SeSame’s^59^ linear preprocessing pipeline using prepSesme “QGCDPIU”. Wherein, raw IDAT data was processed by performing quality masking, background subtraction, dye-bias correction, and detection p-value masking. Then, to ensure we only used informative sites, we estimated per site its Median Absolute Deviation (MAD), retaining only probes with a *MAD >* 0.005. Afterwards, we used minfi’s^60^ “estimateCellCounts2” command together with FlowSorted.Blood.EPIC^61^ reference dataset to estimate the cell type proportions from the bulk sample. We then normalized every sample to have its cell type proportion estimates sum to 1. We then employed TCA on the methylation betas to receive cell type de-convolved methylation profiles. To ensure each site is reliable within each of the cell type specific profiles, we again applied a *MAD >* 0.005 filter. To ensure the data maintains normality assumptions, we employed the RNOmni package^62^ to rank-based inverse normal transform the probe data across samples. Then, similarly to what we did in the transcriptome pre-processing, we performed PCA, and approximated the optimal number of principal components as covariances by using the Gavish-Donoho method^58^. Methylomics were done on blood leukocytes.

### 4.8 Genotype-by-Environment Interaction Mapping

To identify *G × E* interactions, we performed *cis*-association mapping using the cis_ nominal mode of tensorQTL together with the interaction parameter^63^. Specifically, we used similar tensorQTL parameters to those used previously^12,63–65^. These were chosen as they account for the genomic regulatory architectures of the transcriptome and methylome, while maintaining computational viability and were grounded in biological reasoning. The *cis*-window sizes were defined as 1 Mb for eQTLs and 5 kb for meQTLs respectively, and genetic variants were filtered to those with a minor allele frequency (MAF) *>* 0.05. Significance threshold is set to the standard cis-QTL threshold of *p.value <* 1 *∗* 10*^−5^*. The interaction model was defined as:

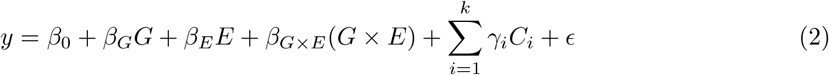

where *y* represents the normalized molecular phenotype, either gene expression or methylation levels, for a gene or a CpG methylation site, respectively. *G* is the genotype dosage, and *E* represents the continuous interaction variable, which is a metabolic or proteomic module retrieved from WGCNA as described above. The model incorporates *k* covariates (*C_i_*), representing the ancestry and latent molecular principal components, the number of which was determined via the Gavish-Donoho method^58^, with *ɛ* representing the residual error. By nature, this analysis also identified variants with a main effect, as well as environmental effect.

### 4.9 GxE Discovery Determinants

To map the biological associations rules determining the number of discovered *G × E* interactions across different molecular layers, we modeled the total count of significant *G × E* hits (unique eGene or methylation site), using a negative binomial generalized linear model (GLM)^66^. The model was formulated as follows:

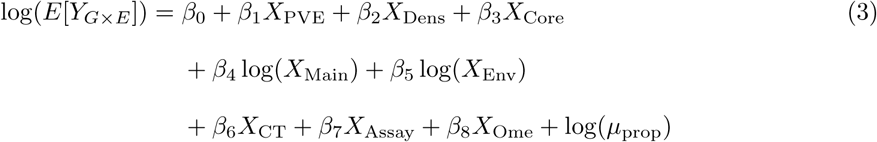

where *Y_G×E_* equals the number of *G × E* hits discovered per module in an assay, in a cell type, *X_P V E_* represents the percentage of variance explained by the module eigengene, *X_Dens_* is the module density, *X_Core_* is the network coreness, *X_Main_* and *X_Env_* are the counts of significant genetic main effects and environmental effects respectively in the same cell type and assay, *X_CT_* is the cell type, *X_Assay_* is the experimental assay, *X_Ome_* is the specific omics layer, and log(*µ_prop_*) is an offset term equal to the log-transformed median proportion of a given cell type within the cohort’s samples.

The inclusion of the offset term log(*µ_prop_*) is to account for varying cellular proportions across the cohort, helping address the difference in discovery power between different cell types. Nonetheless, it is important to note that intra-cell type heterogeneity might further drive differences in cell type *G × E* discovery power, which were not modeled here. Count predictor variables, in particular the number of main genetic effects and number of environmental hits, were log-transformed because their distributions are highly skewed and a transformed scale was considered more appropriate for modeling.

To further map the rules governing the discovery of *G×E*, we performed variance decomposition based on the model deviance. Specifically, We calculated a Pseudo *R*^2^ by:

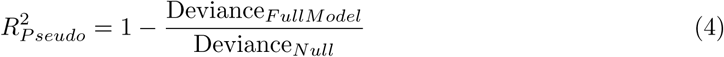

Variable contributions to the total variance were determined using ANOVA. The *R*^2^ contribution for each term was defined as the proportion of the null deviance reduced by each specific variable.

### 4.10 Understanding Molecular Phenotypes with multiple *G × E* hits

We sought to characterize the relationship between the number of proteomic and metabolic MEMs exhibiting molecular phenotypes with significant G×E interactions and the number of cell types in which these interactions were observed. To do so, we fit the following GLM^66^ model:

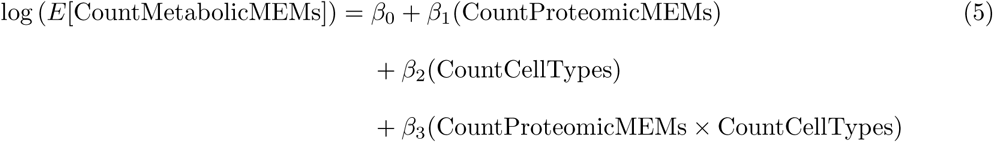

where CountMetabolicMEMs and CountProteomicMEMs represent, respectively, the number of metabolic and proteomic MEMs for which the same molecular phenotype showed a *G×E* interaction, and CountCellTypes represents the number of unique cell types in whom a *G × E* interaction was observed.

### 4.11 Zero-Shot Cross-Cell-Type Prediction of *G × E* Interactions

To examine whether we could predict, using information we have in multiple cell types, whether a molecular phenotype (gene expression or methylation) will be affected by a *G × E* interaction, in a held-out cell type we developed a model. Specifically, using a leave-one-cell-type-out cross-validation (LOCTO) framework, we trained on *G × E* interaction data as well as appropriate molecular phenotype data from the remaining cell types and evaluated the performance on the left-out cell type in a zero-shot frame work, when cell type molecular phenotype is given. During prediction, the model has molecular phenotype data for the particular cell type in which it performs the *G × E* interaction prediction. With the expansion of next generation sequencing and the advent of single cell sequencing, most cell types are molecularly phenotyped^30,67,68^, thus, this prior information is readily available and applicable in zero-shot applications. The model used features from 3 sources: (1) cell-type-specific molecular phenotype features, (2) MEM specificity, and (3) features describing Gene’s *G × E* interaction presence in other cell types. All training-derived statistics were computed inside each fold without the left-out cell type to prevent data leakage from the held-out cell type into the trained model. Specifically, the variables used by the model were: “gene_ resp_rank” which is calculated by ranking all the genes by the total number of MEMs with whom they are associated, and then dividing by the total number of genes, to get a percentile, “log module potency” which is calculated via the log1p of the total number of MEMs with whom the molecular pheno-type is associated with, “gene_n_modules” which is calculated via calculating the number of unique MEMs with whom the molecular phenotype is associated, “module_n_genes” which is calculated via the number of unique molecular phenotypes associated with the MEM, “gene_n_cells” which is calculated via the count of distinct cell types in whom we see a *G × E* interaction with the same molecular phenotype, “expr_x_potency” which is calculated via an interaction term between log expr and log module potency, “log_expr” which is calculated via a log1p of the de-convolution result, “is expressed” which is calculated via classifying above or below 0 of the raw de-convolution output, and “module n cells” which is calculated via the number of cell types in whom the MEM shows a *G × E* interaction affecting the levels of the molecular phenotype.

Classification was performed using XGBoosted trees^69^, we accounted for training class imbalance via inverse frequency weighting correction. The test cell type was never observed during training or model selection, preventing data leakage. we evaluated the models using the area under the precision-recall curve (AUPRC) as our primary measurement. Given the high class imbalance, and AUPRCs dependence on prevalence, we reported the lift over the random baseline (AUPRC / prevalence).

### 4.12 Colocalizing Gene by Environment Interactions with Traits

To identify loci where *G×E* interactions may share a causal variant with a complex trait GWAS, we performed colocalization analysis following a similar approach to that used by Kasela et al^12^. Due to computational constraints, we restricted this analysis to *G× E* results from the first 18 metabolomic molecular exposure modules. In particular, we applied the `coloc.abf` method as implemented in the `coloc R` package^70^, restricting analyses to *G × E* loci where the lead interaction variant reached *p*_int_ *<* 10*^−^*^5^. Variants within a 200 kb window of the lead interaction variant were clumped together to define each locus. We required a minimum of 50 variants overlapping between the *G×E* locus and the GWAS summary statistics per locus, to perform the colocalization analysis. The MHC region was excluded. Prior probabilities were set as *p*_1_ = 10*^−^*^4^, *p*_2_ = 10*^−^*^4^, and *p*_12_ = 5 *×* 10*^−^*^6^, and a posterior probability of a shared causal variant (*PP.H*_4_ *>* 0.50) was used to define colocalization^12^. A subset of colocalized loci were additionally examined manually by visual inspection. The analysis utilized a set of harmonized and curated GWAS^20^.

## 5 Acknowledgments

We thank Andrew Fire for help with advising and feedback. We thank members of Stephen Montgomery’s and Andrew Fire’s laboratories for general guidance and feedback on this work.

We acknowledge and thank the researchers and participants who provided biological samples and data for TOPMed, and in particular MESA.

Furthermore, Whole genome sequencing (WGS) for the Trans-Omics in Precision Medicine (TOPMed) program was supported by the National Heart, Lung and Blood Institute (NHLBI). WGS for “NHLBI TOPMed: Multi-Ethnic Study of Atherosclerosis (MESA)” (phs001416.v1.p1) was performed at the Broad Institute of MIT and Harvard (3U54HG003067-13S1). Centralized read mapping and genotype calling, along with variant quality metrics and filtering, were provided by the TOPMed Informatics Research Center (3R01HL-117626-02S1). Phenotype harmonization, data management, sample-identity QC, and general study coordination were provided by the TOPMed Data Coordinating Center (3R01HL-120393-02S1). The Multi-Ethnic Study of Atherosclerosis (MESA) projects are conducted and supported by the National Heart, Lung, and Blood Institute (NHLBI) in collaboration with MESA investigators. Support for MESA is provided by contracts 75N92025D00022, 75N92020D00001, HHSN268201500003I, N01-HC-95159, 75N92025D00026, 75N92020D00005, N01-HC-95160, 75N92020D00002, N01-HC-95161, 75N92025D00024, 75N92020D00003, N01-HC-95162, 75N92025D00027, 75N92020D00006, N01-HC-95163, 75N92025D00025, 75N92020D00004, N01-HC-95164, 75N92025D00028, 75N92020D00007, N01-HC-95165, N01-HC-95166, N01-HC-95167, N01-HC-95168, N01-HC-95169, UL1-TR-000040, UL1-TR-001079, UL1-TR-001420, UL1TR001881, and R01HL105756. The authors thank the MESA participants and the MESA investigators and staff for their valuable contributions. A full list of participating MESA investigators and institutions can be found at http://www.mesa-nhlbi.org

## Declarations

### 5.3 Ethics approval and consent to participate

This research is IRB exempt and is on anonymized summary data.

### 5.4 Data availability

TOPMed MESA data was accessed via controlled access permission, and can be downloaded via the database of Genotypes and Phenotypes (dbGaP) through study accession number phs001416.v4.p1 (https://www.ncbi.nlm.nih.gov/gap).

### 5.5 Code availability

Code used in this work is available via Github at https://github.com/haimkru/Celluar-gene-by-molecular-exposure-analysis.

## Supplementary information

Supplementary Tables 1–4 are found in Zenodo 10.5281/zenodo. 22865894, as well as significant (*p.value <* 1 *∗* 10*^−^*5) per cell type *G × E* interactions.

## Extended Data Figures

**Extended Data Fig. 1:**
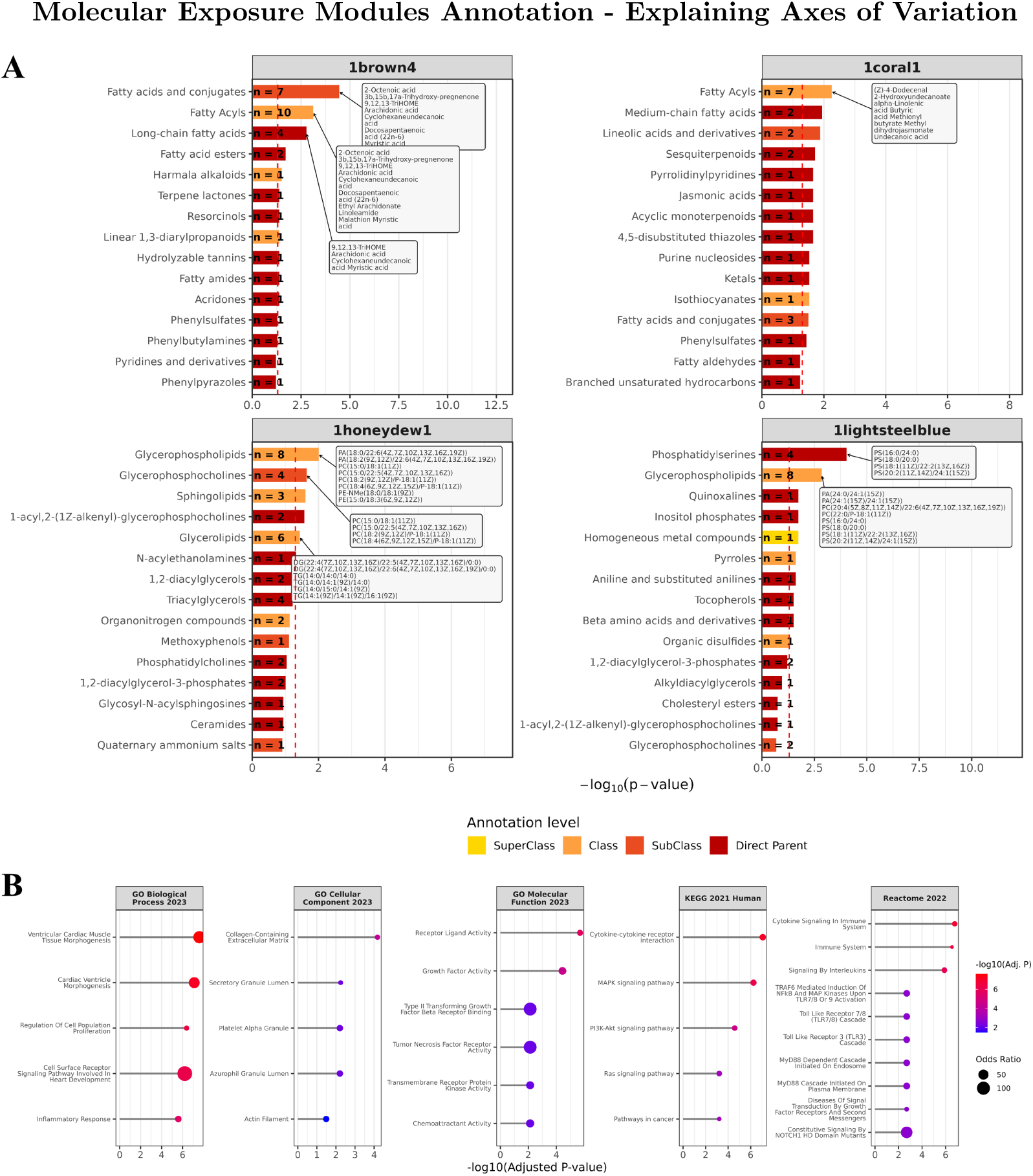
To shed some light into what composes the Molecular Exposure Modules, we chose to perform an over representation analysis(ORA) on the metabolites found in the metabolic MEMs, and on the proteins found in the proteomic module (see Methods 4.4). **a** To annotate metabolites, we performed ORA against HMDB’s chemical structure terms. Plotted are the metabolome features who are highly explainable. For a full list, see supplementary table 1. **b** To annotate proteins, we performed ORA using enrichR against the GO, KEGG, and Reactome databases. Plotted is proteomic MEM yellow, associated with BMI,CRP, and inflammation. For results across all proteomic MEMs, see supplementary table 2.

**Extended Data Fig. 2:**
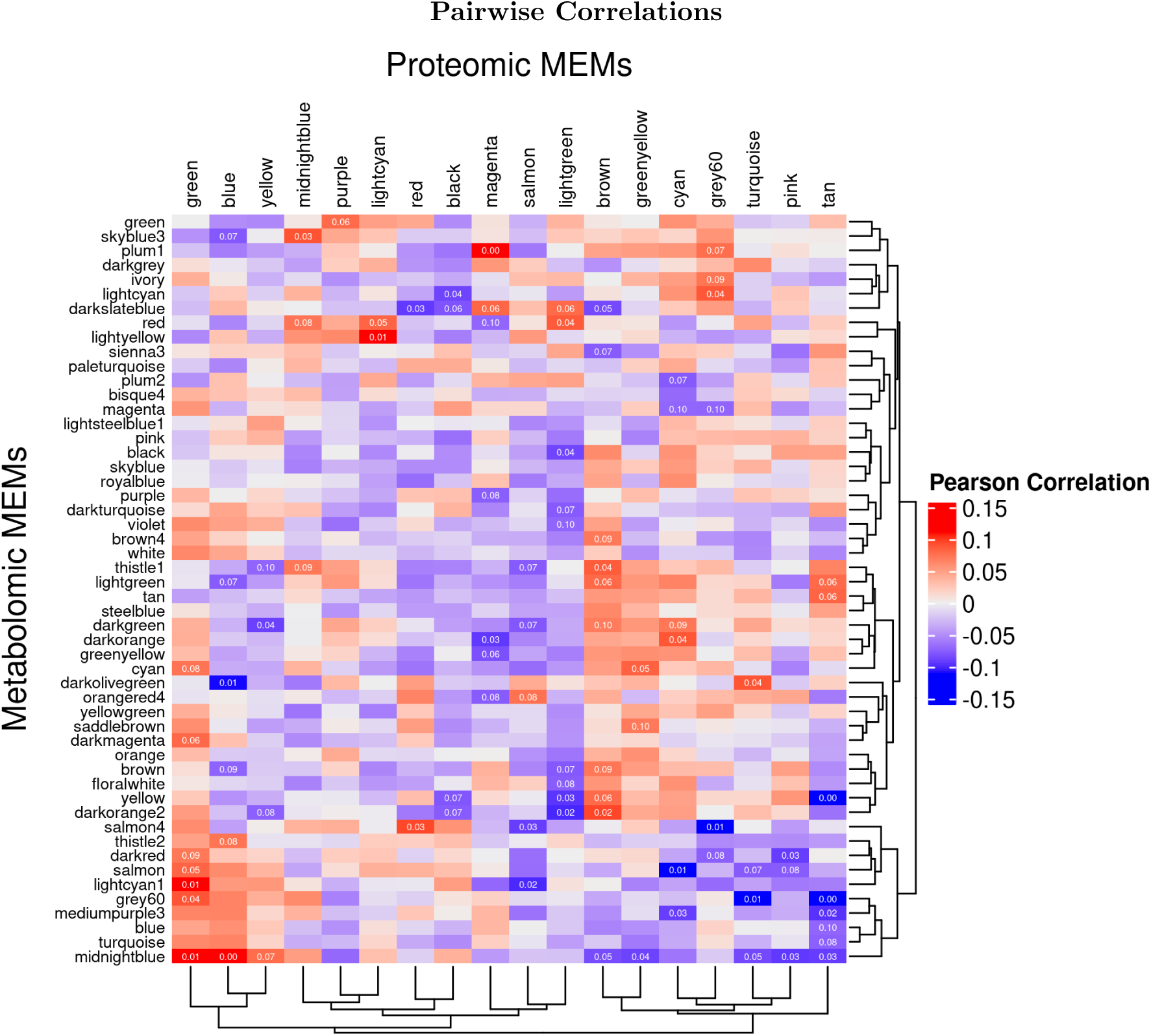
Pairwise correlations between proteomic and metabolic MEMs.

**Extended Data Fig. 3:**
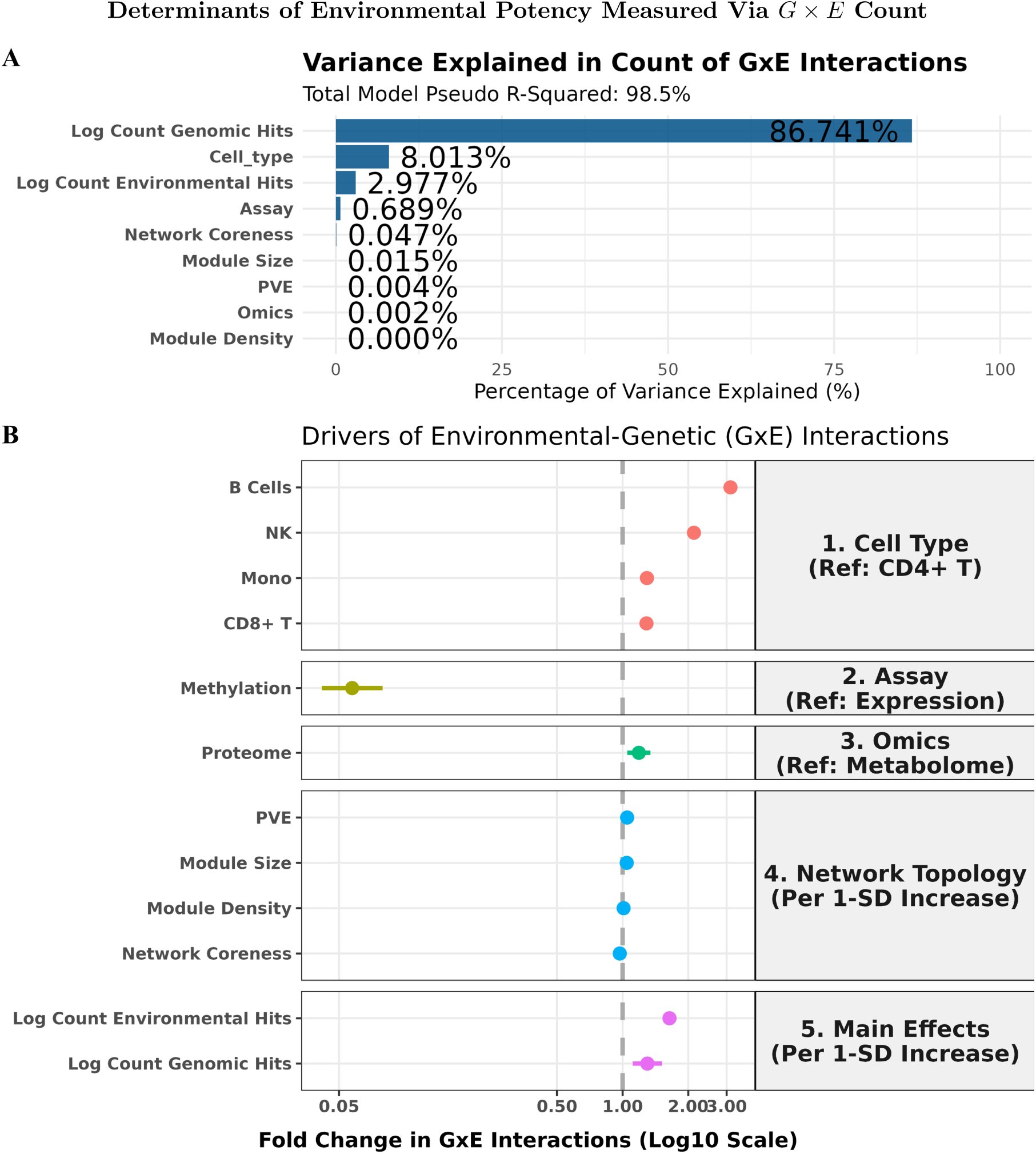
To better understand the rules that govern the discovery count of unique molecular phenotypes for whom a *G E* interaction is detected, with used a multivariate model. **A**, Variance explained by each determinant of *G E* interaction count. Count of main effect variants is the variable with the highest explainability, followed by cell type. **B**, Effect size of each determinant of count *G E* interaction count per cell type-MEM-ome-assay combo. B cells exhibit an especially high rate of discovery. *For detailed outline of method, see Methods 4.9.

**Extended Data Fig. 4:**
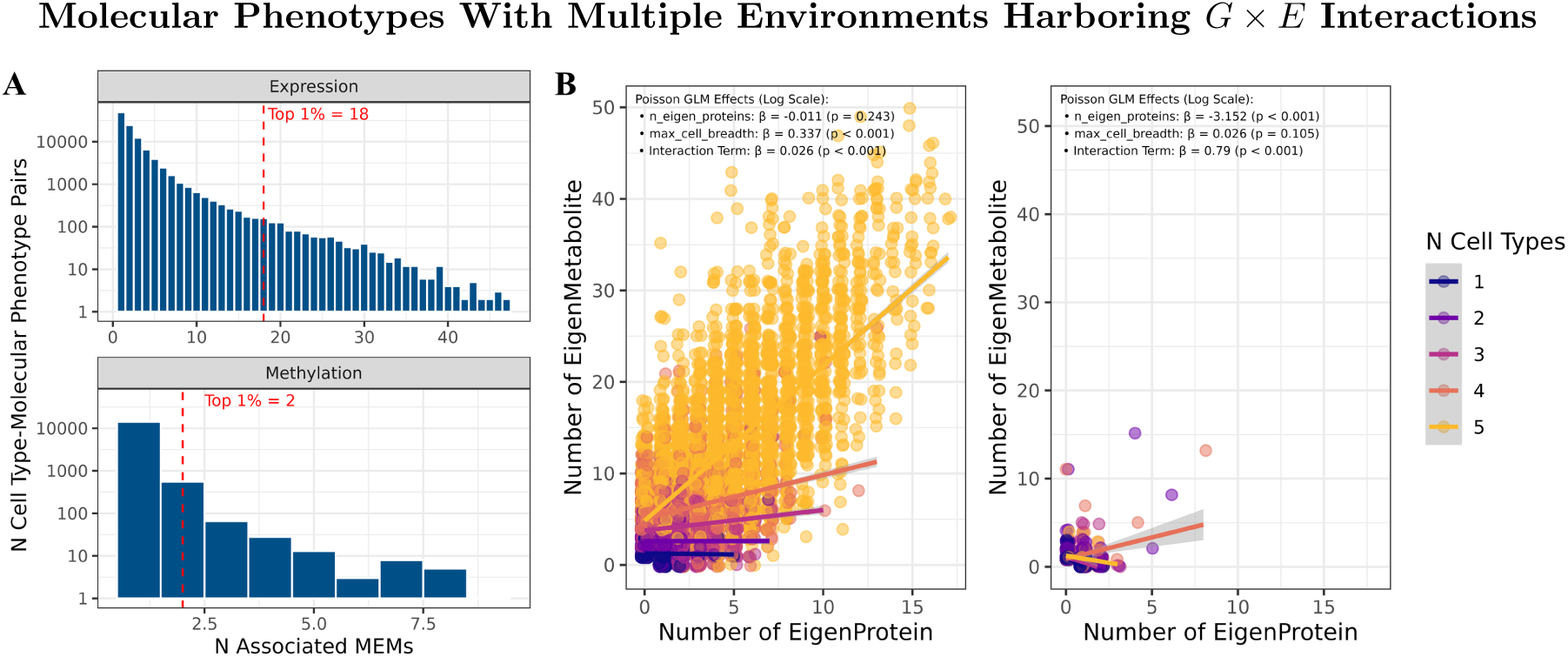
**A**, Distribution, of per each molecular phenotype (unique eGene or CpG Methylation Site) the number of MEMs with whom we discovered *G* X *E* association. **B**, Each dot represents a Molecular phenotype, with the x axis representing the number of associated proteomic MEMs, and the Y axis representing the number of Metabolic MEMs the molecular phenotype is associated with. The coloring is by the number of unique associated cell types. P. values are from the GLM interaction models, see Methods 4.10.

**Extended Data Fig. 5:**
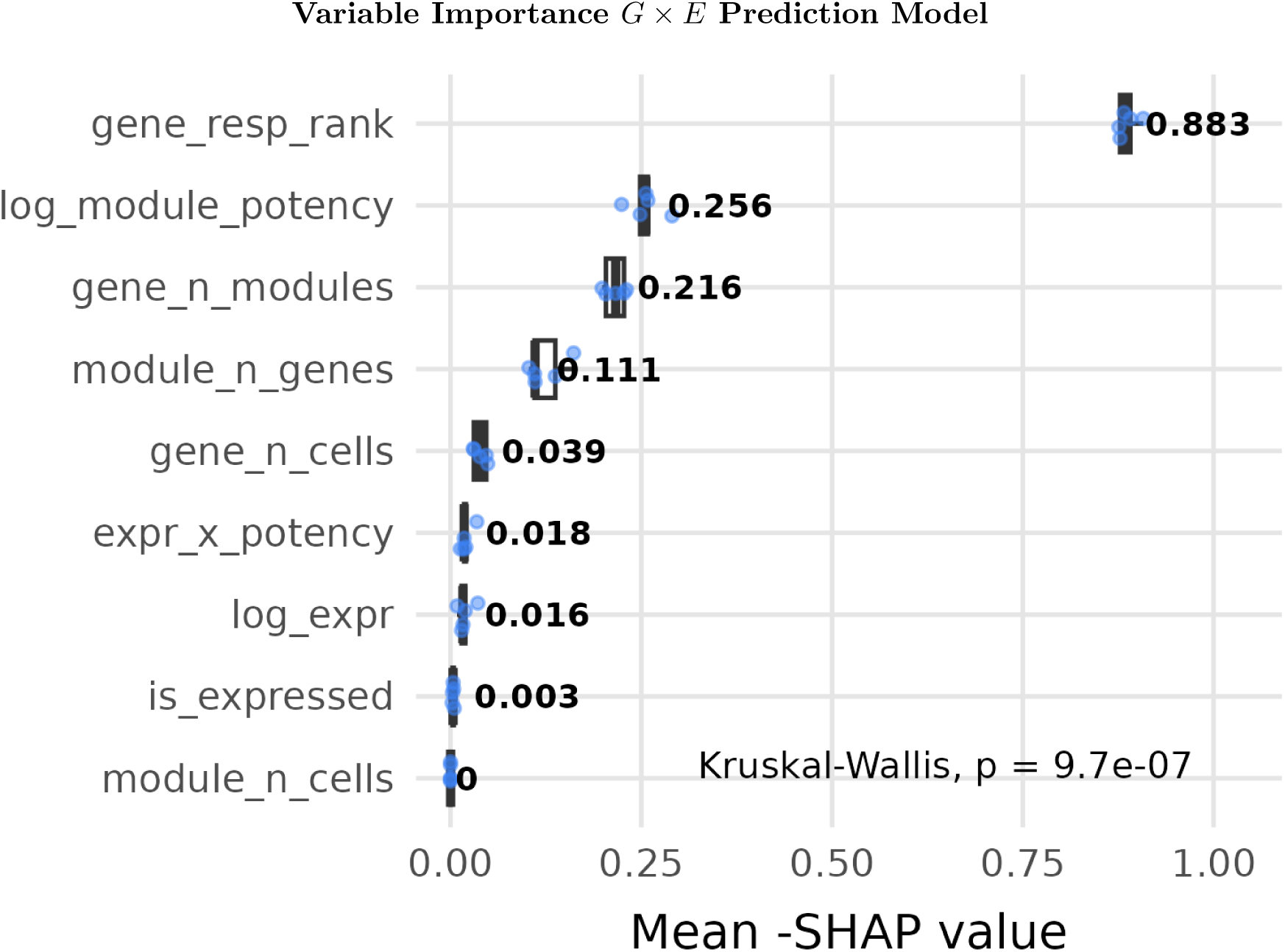
To better understand the inner workings of the *G* X *E* eGene-MEM prediction zero shot transfer learning XGBoost tree model, we used SHAP^19^ to determine variable importance.

**Extended Data Fig. 6:**
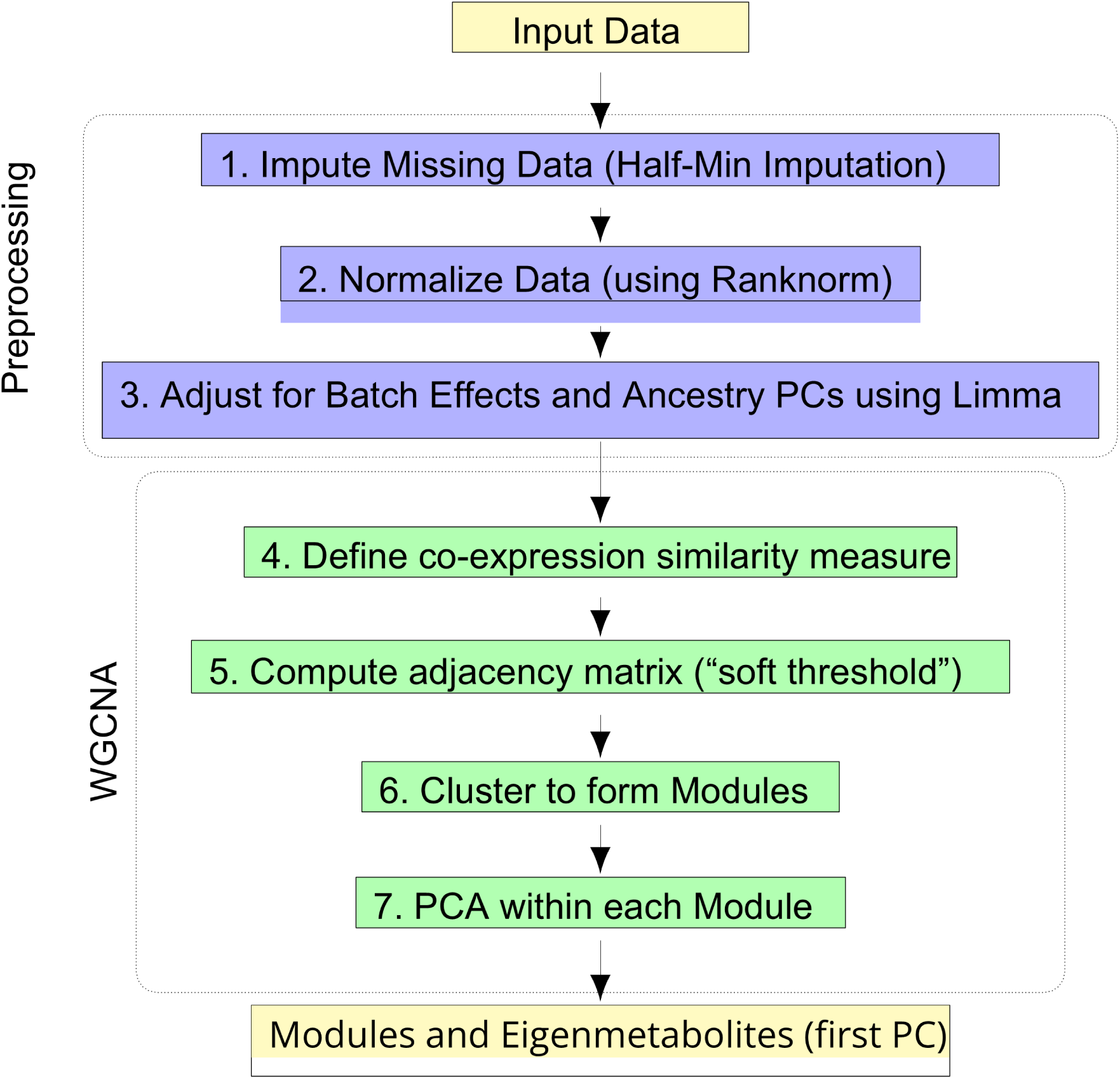
For the pre processing steps, we impute the missing data to enable down-stream analyses, normalize the data to ensure that the expression values are comparable across samples and adjust for batch effects and Ancestry PCs to account for these potential confounding factors. VST normalization gave the best results. (ranknorm, VST and invnorm were explored).

**Extended Data Fig. 7:**
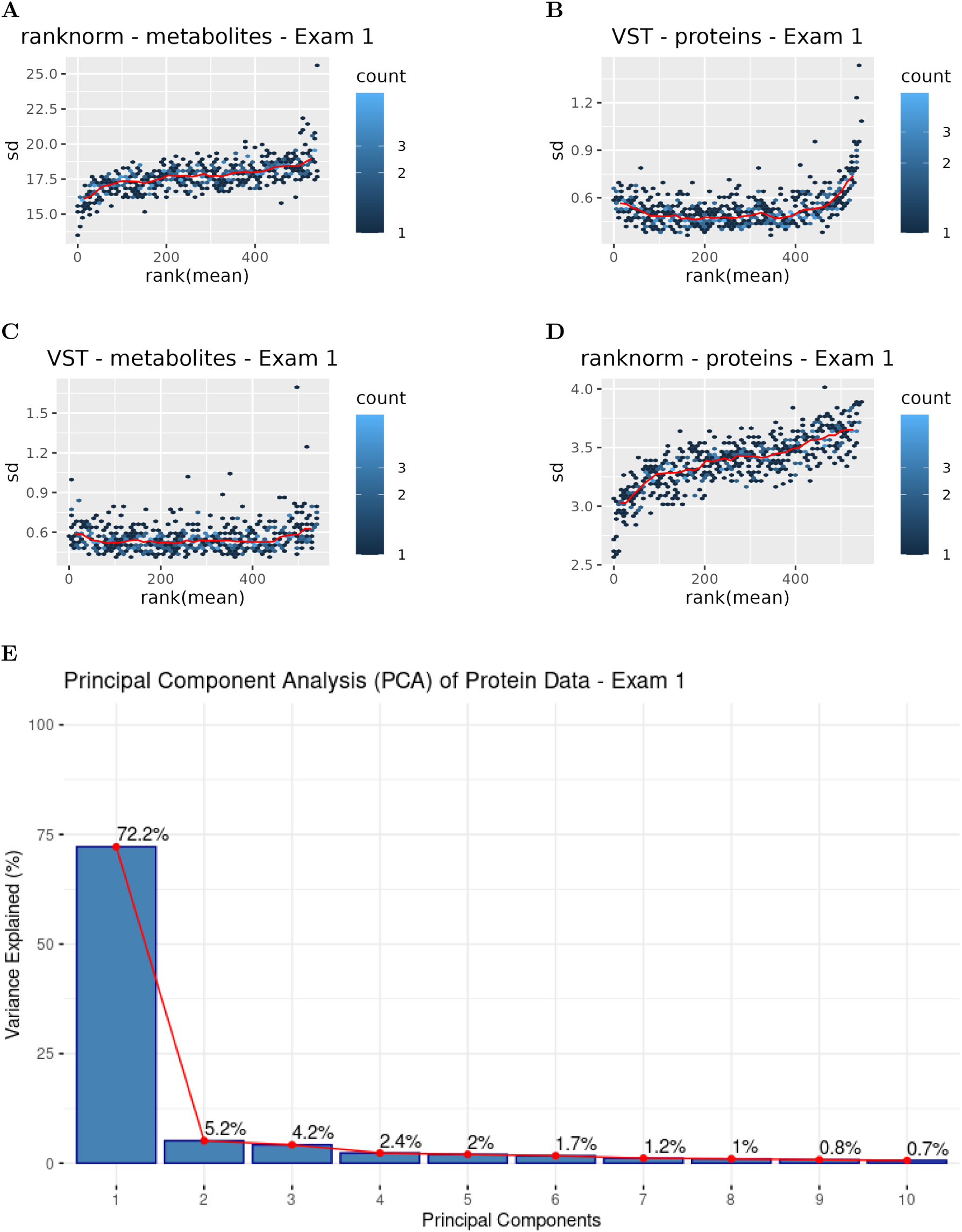
**A**, Mean–variance relationship for plasma metabolites (n = 6,042) at Exam 1 under rank-based inverse normal transformation (Ranknorm). While Ranknorm reduces the influence of outliers, substantial variability remains across features. **B**, Mean–variance relationship for plasma proteins (n = 1,317) at Exam 1 under Ranknorm normalization. Residual heteroscedasticity is observed across protein features. **C**, Mean–variance relationship for plasma metabolites (n = 6,042) at Exam 1 after variance stabilizing transformation (VST). VST improves variance stabilization across the dynamic range of metabolite abundances compared to Ranknorm. **D**, Mean–variance relationship for plasma proteins (n = 1,317) at Exam 1 following VST normalization. Variance is more consistent across the range of protein expression values. **E**, Principal Component Analysis (PCA) of Protein Data.

